# Specific pattern of melanin-concentrating hormone (MCH) neuron degeneration in Alzheimer’s disease and possible clinical implications

**DOI:** 10.1101/2021.01.27.21250608

**Authors:** Mihovil Mladinov, Jun Yeop Oh, Cathrine Petersen, Rana Eser, Song Hua Li, Panos Theofilas, Salvatore Spina, William W. Seeley, Jackson C. Bittencourt, Thomas C. Neylan, Lea T. Grinberg

## Abstract

**Study Objectives:** The lateral hypothalamic area (LHA) is one of the key regions orchestrating sleep and wake control. It is the site of wake-promoting orexinergic and sleep-promoting melanin-concentrating hormone (MCH) neurons, which share a close anatomical and functional relation. The aim of the study was to investigate the degeneration of MCH neurons in Alzheimer’s disease (AD) and progressive supranuclear palsy (PSP), and relate the new findings to our previously reported pattern of degeneration of wake-promoting orexinergic neurons

**Methods:** Post-mortem human brain tissue of subjects with AD, PSP and controls was examined using unbiased stereology. Double immunohistochemistry with MCH- and tau-antibodies on formalin-fixed, celloidin embedded tissue was performed.

**Results:** There was no difference in the total number of MCH neurons between AD, PSP and controls, but a significant loss of non-MCH neurons in AD patients (p=0.019). The proportion of MCH neurons was significantly *higher* in AD (p=0.0047). No such a difference was found in PSP. In PSP, but not AD, the proportion of tau+ MCH neurons was *lower* than the proportion of tau+ non-MCH neurons (p=0.002). When comparing AD to PSP, the proportion of tau+MCH neurons was higher in AD (p<0.001).

**Conclusions:** MCH neurons are more vulnerable to AD than PSP pathology. High burden of tau-inclusions, but comparably milder loss of MCH neurons in AD, together with previously reported orexinergic neuronal loss may lead to a hyperexcitability of the MCH system in AD, contributing to wake-sleep disorders in AD. Further experimental research is needed to understand why MCH neurons are more resistant to tau-toxicity compared to orexinergic neurons.

**STATEMENT OF SIGNIFICANCE:** This is the first study to investigate the involvement of melanin-concentrating hormone (MCH) neurons in patients with Alzheimer’s disease and progressive supranuclear palsy. MCH neurons are key regulators of sleep and metabolic functions, and one of the major neuronal populations of the lateral hypothalamic area (LHA), but still underexplored in humans. Uncovering the pathology of this neuronal population in neurodegenerative disorders will improve our understanding of the complex neurobiology of the LHA and the interaction between MCH and orexinergic neurons. This new knowledge may open new strategies for treatment interventions. Further, this study represents a fundament for future research on MCH neurons and the LHA in tauopathies.

## INTRODUCTION

Tauopathies, neurodegenerative conditions with abnormal tau protein deposits, feature a progressive decline of cognitive functions. Confluent data show that sleep disturbances are also highly prevalent and may manifest early in the course of tauopathies, sometimes even preceding the cognitive symptoms.^1–4^ Sleep disturbances impairs the quality of life of patients and caregivers alike, leading to increased institutionalization.^5–7^ The neurobiological basis of sleep disturbances in tauopathies remains unclear, and generalized treatment usually used for sleep disturbances, such as benzodiazepines and Z-drugs, are not recommended due to their risks and side effects.^8,9^ Thus, there is an urgency to clarifying the neurobiological underpinning of the various sleep dysfunction patterns in seen neurodegenerative diseases to enable effective symptomatic and disease-modifying treatment^10^.

The lateral hypothalamic area (LHA) is a key brain region orchestrating sleep and wake control. Together with noradrenergic neurons of the locus coeruleus and histaminergic neurons of the tuberomammillary nucleus, orexinergic neurons promote arousal. For instance, loss of orexinergic neurons underlies narcolepsy ^11^. In a recent study, we showed a profound loss of orexinergic neurons in Alzheimer’s disease (AD), but a relative sparing of the same neurons in 4-repeat tauopathies, ^12^ providing a direct neuropathological explanation for the disruption of vigilance and excessive daytime sleepiness observed in AD, but not in 4-repat tauopathies.^13–15^. Loss of orexinergic neurons were also reported in Parkinson’s disease^16^ and Huntington’s disease.^17^

Besides orexinergic neurons, the LHA houses other important neuronal populations, mainly melanin-concentrating hormone (MCH) neurons. MCH neurons have a close anatomical and functional relation to orexinergic neurons, and both neuropal populations intermingle in LHA (**Fig 1**). Although, MCH neurons are poorly defined anatomically, their highest density is in the LHA, in an area delimited by the dorsal fornix, mamillotegmental and mamillotalamic tracts, and the third ventricle.^18–20^ Whereas orexinergic neurons promote arousal, MCH neurons mainly promote and help to retain the REM-phase of sleep in animal models,^21–23^ thus contrabalancing the orexinergic effects. MCH neurons also regulate motivated and feeding behavior^24,25^ and promote positive energy balance.^26–28^ Thus, MHC neurons are essential for coordinating sleep with other vital functions and integrating these functions into a 24-hour circadian rhythm. Despite their physiological importance from fish to mammals, MCH neurons have barely been characterized in humans and changes in these neurons are yet to be characterized in neurodegenerative diseases.

**Figure 1.**
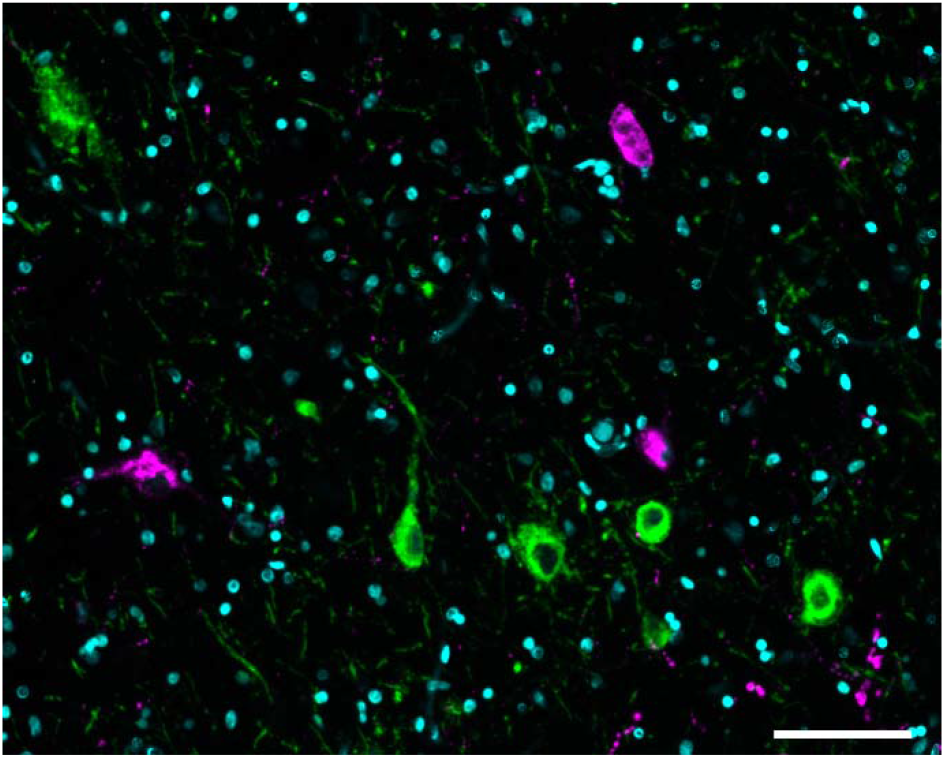
Neurons in the lateral hypothalamic area (LHA) do not co-express MCH and orexin. The figure is a representative image of double-staining immunohistochemistry for MCH (green) and orexin (magenta). The picture was taken from a control subject. Scalebar 100um

Here, we provide a comprehensive and quantitative morphological study of MCH neurons in human brains with AD, progressive supranuclear palsy (PSP, a 4-repeat tauopathy featuring hyperinsomnia and preservation of orexinergic neurons), and normal controls. We used unbiased stereology and double labeled immunohistochemistry to quantify the number of MCH neurons, changes in the proportion of LHA neurons in AD and PSP and burden of tau-pathology. Finally, we related the new findings to our previously reported pattern of degeneration of wake-promoting orexinergic neurons. In addition to unveiling new treatment targets for tauopathies, this study leverages neurodegenerative diseases as natural lesions to investigate the neurobiology of sleep in humans.

## METHODS

### Participants and neuropathological diagnosis

The research was approved by the IRB at UCSF. Disease subjects were selected based on a primary neuropathological diagnosis of AD, or PSP and absence of any other significant neurodegenerative or cerebrovascular changes. Normal control subjects (NC) were free of any cognitive impairment (CDR□=□0); neurological or neuropathological diagnosis. PSP, and NC subjects scored as A≤1B≤2C0 according to the NIA-AA guidelines for the neuropathological assessment of AD^29^. Tissues along with extensive clinical and neuropathological records were sourced from the Neurodegenerative Disease Brain Bank (NDBB) from the University of California, San Francisco (UCSF) and Brazilian BioBank for Aging Studies (BBAS) from the University of Sao Paulo^30^ (Table 1). The NDBB receives brains from patients seen at the UCSF Memory and Aging Center. BBAS is population-based and hosts a high percentage of NC that are not available in NDBB. Neuropathological assessments were performed using standardized protocols and followed the internationally accepted criteria for neurodegenerative diseases^31–33^. Demographic data of included subjects are summarized in **Table 1**.

**Table 1.** Demographics of the examined subjects

|  | AD | PSP | controls |
| --- | --- | --- | --- |
| N total (female/male) | 10 (4/6) | 10 (2/8) | 5 (3/2) |
| N Asian/Black/Caucasian/multiple | 0/0/10/0 | 1/0/8/1 | 0/1/4/0 |
| Age at death (years), mean (SD) | 62.8 (3.94) | 71.2 (8.84) | 74.8 (13.66) |
| Disease duration (years), mean (range) | 10.4 (6-17) | 8.2 (3-13) | 0 (0) |
| CDR score, median (range) | 3 (1-3) | 1.5 (0.5-2) | 0 (0) |
| Braak stage, median (range) | 6 (6) | 2 (1-4) | 0 (0-2) |
| PMI (hours), mean (SD) | 11.5 (10.1) | 15.4 (23.5) | 12.2 (3.3) |

AD, Alzheimer's disease; CDR, clinical dementia rating scale; N, number of examined subjects; PMI, postmortem interval; PSP, progressive supranuclear palsy; SD, standard deviation.

### Tissue processing and immunohistochemistry

The region of interest containing MCH neurons was identified using the Atlas of the Human Brain,^34^ the BrainSpan Atlas of the Developing Human Brain,^35^ and previous publications on anatomy of MCH neurons.^18–20,36^ Hypothalamic blocks containing the whole region of interest (ROI) were formalin-fixed and celloidin embedded,^37^ and cut in serial 30-µm thick coronal or horizontal sections. Section orientation does not affect the optical fractionator probe in stereology. The specific protocol for tissue processing has been previously described (see ^38^ and Supplementary material). Histological section underwent double-labeling immunohistochemistry using free floating methods. After inactivation of endogenous peroxidase (0.3% H202 in methanol), antigen retrieval (0.01 M citrate buffer, 0.05% Tween-20, pH 6.0 in PBS at 95.7°C for 50 min) and incubation in blocking buffer, sections were double-stained with mouse monoclonal CP13 antibody for phospho-Ser202 tau (1:1000, kind gift of Peter Davies, NY) and rabbit polyclonal anti-MCH serum (1:1000, kind gift of Joan Vaughan, Salk Institute, CA) overnight at room temperature. Subsequently, sections were incubated in secondary biotinylated anti-rabbit (1:400, Vector Labs) and secondary conjugated-HRP anti-mouse antibodies (1:400, Advansta) for 1 hour at room temperature. The CP13- and MCH-stainings were developed using immPACT DAB Peroxidase (HRP) Substrate Kit (SK4105, Vector Labs) and Vectastain ABC-AP kit (AK-5000, Vector Labs), respectively. In order to estimate the total number of neurons all sections were counterstained using gallocyanin as a nucleic acid stain at a previously optimized pH (pH 1.9 − 2.1).

To rule out possible co-expression of MCH and orexin, we performed double immunohistochemistry, of MCH and orexin (Orexin A, rabbit polyclonal, H-003-30, 1:500, Phoenix Pharmaceuticals). A representative staining showing absebce of co-expression of this two markers is depicted in **Figure 1**.

### Stereological analysis

The stereological analyses were performed using the StereoInvestigator v.10 software (MBF Bioscience, Williston, VT, USA) installed on a Zeiss Imager A2 microscope. Counting was made for MCH-positive (MCH+), tau-positive (Tau+), MCH and tau-positive (MCH+/Tau+), and double negative neurons using the optical fractionator probe.^39^ **Figure 2** depicts a representative staining of these four classes of neurons. The ROI was delineated at 5x, and the neuronal counting was performed using a 63x objective. The stereological parameters were determined using the “resample–oversample” analysis probes included in the StereoInvestigator software. The guard zone was set at 5 µm and the dissector height at 12.5 µm. The coefficient of error (CE) range was calculated following the methods of Gundersen and Schmitz-Hof.^40,41^

**Figure 2.**
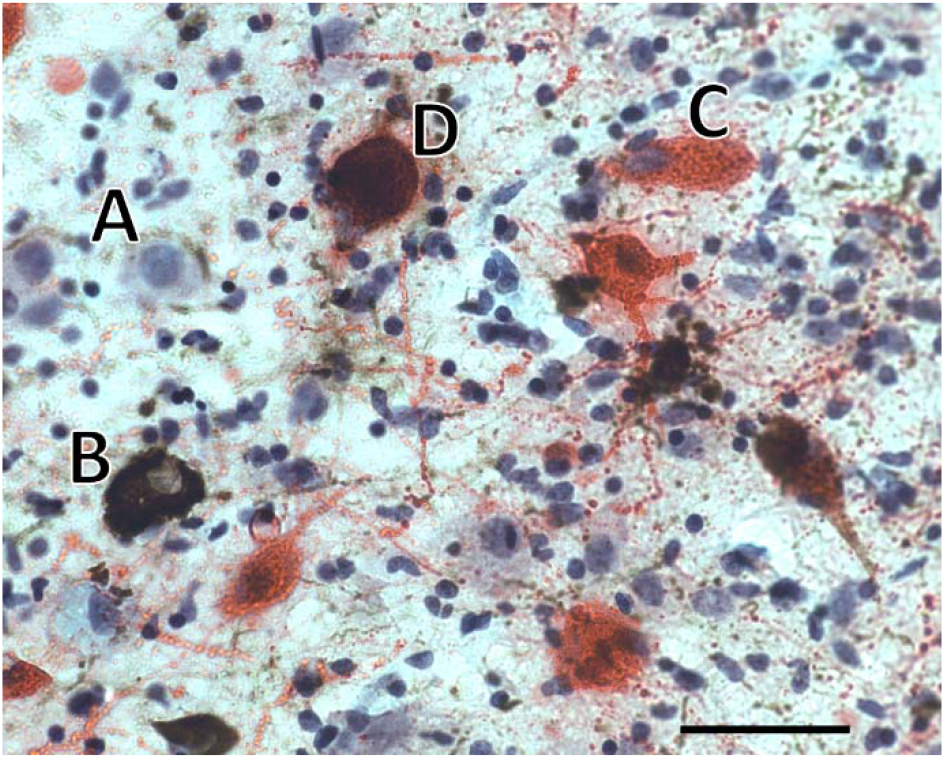
Neuronal classification using double-staining immunohistochemistry. (A) a non-MCH neuron without tau inclusion (light blue), (B) a neuron with phospho-tau inclusion (brown); (C) a neuron positive for MCH only (red); (D) a neuron positive for both phospho-tau and MCH. This photo was taken from the lateral hypothalamic area from a case with Alzheimer’s disease. Scalebar 50um

### Statistical analyses

Statistical analyses were performed using R Statistical Software (version 3.6.1; R Foundation for Statistical Computing, Vienna, Austria). The proportion of neurons positive for MCH was determined by dividing the number of neurons positive for MCH by the total neuron population, represented as a percent. Two-tailed non-parametric Wilcoxon signed rank tests were used for comparisons of MCH- and non-MCH neuronal numbers, and proportions of total neurons within each group (AD, PSP, control). For comparison of neuronal numbers and their proportions between groups, two-tailed non-parametric Mann-Whitney U tests were used. Multiple linear regression was used to correct for age, sex, and Braak stage.

## RESULTS

**Table 1** summarizes demographic, neuropathological and clinical data of the subjects. The clinical severity of neurodegenerative conditions correlates with neuronal loss. Hence, we first quantified the total number of neurons (with and without tau-inclusions) in the ROI. There was no difference in the total number of MCH neurons between AD, PSP and controls, but a significant loss of non-MCH neurons in AD patients. In the AD group, we observed 49.4% fewer non-MCH neurons compared to normal controls (p=0.0193). whereas, the lower number of MCH neurons observed did not reach statistical significance (p=0.0992). In PSP, neither the number of non-MCH nor MCH neurons were significantly different compared to controls. Although the proportion of MCH neurons in the ROI was similar in all groups, subjects with AD had a significantly higher proportion of MCH neurons compared to controls (25.65% vs 21.52%, p=0.0047). Detailed data is provided in **Table 2**.

**Table 2.** Comparison of estimated mean numbers and proportions of neurons in the region of interest by group.

|  | Control | AD | p value | PSP | P value |
| --- | --- | --- | --- | --- | --- |
| # non-MCH neurons (SD) | 241382 (71979) | 122193 (77666) | **0.0193** | 154971 (104127) | 0.2544 |
| # MCH neurons (SD) | 65252 (17096) | 41009 (24938) | 0.0992 | 45249 (23985) | 0.1645 |
| % MCH of total neurons (SD) | 21.52% (1.71) | 25.65% (2.56) | **0.0047** | 24.45% (4.36) | 0.2065 |
| % tau+ of non-MCH neurons (SD) | 0.06% (0.04) | 15.94% (3.92) | **0.0007** | 17.39% (5.96) | **0.0007** |
| % tau+ of MCH neurons (SD) | 0.02% (0.03) | 16.24% (4.75) | **0.0026** | 4.85% (2.12) | **0.0026** |

#, number of neurons; %, percentage of neurons; AD, Alzheimer's disease; MCH, melanin-concentrating hormone; p value, compared to controls; PSP, progressive supranuclear palsy; SD, standard deviation

To further investigate the pattern of degeneration and vulnerability of MCH neurons compared to other neuronal populations in the ROI, we analyzed the proportion of neurons with tau-inclusions and related it to the extent of neuronal loss. In AD, both MCH and non-MCH neurons showed a similar tau burden of around 16% (**Table 2**). However, the PSP group showed a significantly lower involvement of MCH neurons (4.58%) compared to non-MCH neurons in the ROI (17.39%, p=0.0020). A comparison between AD and PSP showed that the proportion of tau-positive MCH neurons was significantly higher in AD compared to PSP (16.24% vs 4.85%, p<0.0001). There was no difference in the tau burden of non-MCH neurons in the ROI between AD and PSP (15.94% vs 17.39%, p=0.1230).

## DISCUSSION

This is the first stereological study addressing the fate of MCH neurons in patients with AD and PSP. MCH neurons are evolutionary conserved,^42^ and along with orexinergic neurons they represent the major neuronal population of the lateral hypothalamic area (LHA), an area integrating sleep and metabolic functions.^43^ Our previous study shows that the involvement of orexinergic neurons is a major feature of LHA degeneration in AD and PSP.^12^ Considering the critical role of MCH neurons, we now investigated the impact of AD and PSP pathology in this neuronal population. In contrast to orexinergic neurons, we found that MCH neurons exhibited less degeneration in both AD and PSP. In AD, MCH neurons showed notable tau-burden, but no significant reduction in neuronal number. Compared to AD, subjects with PSP had a lesser impact in MCH neurons, both in terms of tau-inclusions and neuronal number. These findings were unexpected, considering our hypothesis that there would be a major involvement of MCH neurons in PSP. Namely, PSP features protracted insomnia^7,44^ and a reduction in the number and duration of REM-sleep periods,^7,45^ which may indicate a dysfunction of MCH neurons. The PSP group showed no significant loss of MCH neurons in the examined region, with less than 5% of MCH neurons containing tau inclusions (**Table 2**). Regarding our current findings in AD, we found a relatively preserved number of MCH neurons. However, a notable proportion (around 16%) of these neurons did have tau inclusions. As the surrounding neurons of the examined region had the same proportion of tau inclusions, we conclude that MCH neurons show substantial AD-pathology, but are relatively resistant to cell death. The relative sparing of MCH neurons in both AD and PSP rises the question of what makes this particular neuronal population resistant to tau-induced neurotoxicity. MCH and orexin neurons are part of the same functional systems, they anatomically intermingle, and share the same extracellular space. Both MCH and orexin neurons have widespread projections to several subcortical and cortical brain regions.^18,46,47^ A further open question is the functional consequences of this finding. Although relatively mild, the observed pathology may have significant clinical implications. It is known that in the cerebrospinal fluid (CSF) of AD patients, levels of MCH and other neurotransmitters like orexin and norepinephrine are elevated.^48–50^ This seems like a paradoxical finding considering substantial cell loss of orexinergic and noradrenergic neurons in AD. Further, MCH levels in CSF also positively correlate with the disease severity.^48^ Considering that, we hypothesize that tau-inclusions cause a dysfunction of MCH neurons, eventually leading to hyperexcitation with hyperproduction of MCH, and elevated MCH CSF levels in AD. Further, it is important to consider the aforementioned pathology of orexinergic neurons. They inhibit the MCH neurons through activation of GABAergic neurons to promote wakefulness.^43,51^ Hence, the lack of orexinergic input in the frame of AD pathology may also contribute to a hyperfunction of MCH, leading to disorders of sleep and wakefulness. In fact, MCH facilitates entry into REM sleep,^22,23,52,53^ and stimulation of MCH neurons during NREM sleep increases the transition into REM sleep.^22^ Consequently, the hyperexcitability of MCH neurons may facilitate REM sleep at the expence of slow-wave NREM^49,54^, contributing to the reduction of slow-wave NREM in AD. In contrast to AD, PSP features a hyperarousal state, with significantly shorter sleep time, including reduced N2, N3 and REM stages.^7^ This indicates that in PSP the activity of the arousal system prevails over a deficient sleep promoting system. In our previous study, we reported an extensive involvement of orexinergic neurons in PSP, with loss of orexin expression in 56% of neurons, and 51% of tau-positive neurons. In the current study we showed that MCH neurons exhibit a relatively mild pathology. Consequently, it is possible that the involvement of other sleep promoting nuclei, like neurons in the intermediate nucleus, basal forebrain or parafacial zone contribute to the insomnia in PSP.

From the methodological point of view, the strengths of our study are the use of unbiased stereological method, which provides reliable neuronal counting results; and the thoroughly characterized cases. The demographic difference in age and gender of the compared groups (AD, PSP, controls) is a common limitation of neuropathological studies using human brain tissue. Brain samples of clinically and pathologically well characterized characterized subjects are limited, which necessitates compromises regarding sample number and matching controls. The lower age of onset in the examined AD subjects compared to PSP subjects is a consequence of recruitment of early-onset AD patients at our institution. However, we do not expect that the demographic differences introduced a bias concerning the results and conclusions of the study. First, all analyses included a correction for age and sex. Further, the included subjects were in terminal disease phases with fully developed pathology, at which point disease modulating factors like demographics are of less relevance.

In conclusion, this study provides insight about the selective vulnerability of sleep-promoting neurons in AD and PSP, and complements the knowledge regarding the extent and pattern of LHA degeneration in tauopathies. We propose that tau-pathology leads to hyperexcitation of MCH neurons, contributing to dysregulation of vigilance and reduction of NREM sleep in AD. However, what makes the MCH neurons resistant to tau-toxicity in terms of cell death warrants further research on animal models, and thorough phenotypization of specific neuronal populations like single-cell RNA expression analysis. From the clinical point of view, the encouraging finding that MCH neurons in AD show high burden of tau-pathology, but comparably milder cell loss, suggests that future targeted therapies could rescue the function of MCH neurons even in advanced stages of AD.

## Data Availability

All original data will be available upon request

## References

1. Webster L, Costafreda Gonzalez S, Stringer A, et al. Measuring the prevalence of sleep disturbances in people with dementia living in care homes: a systematic review and meta-analysis. Sleep. 2020;43(4).

2. Wennberg AMV, Wu MN, Rosenberg PB, Spira AP. Sleep Disturbance, Cognitive Decline, and Dementia: A Review. Semin Neurol. 2017;37(4):395–406.

3. Hahn EA, Wang HX, Andel R, Fratiglioni L. A change in sleep pattern may predict Alzheimer disease. Am J Geriatr Psychiatry. 2014;22(11):1262–1271.

4. Lim AS, Kowgier M, Yu L, Buchman AS, Bennett DA. Sleep Fragmentation and the Risk of Incident Alzheimer’s Disease and Cognitive Decline in Older Persons. Sleep. 2013;36(7):1027–1032.

5. Spira AP, Kaufmann CN, Kasper JD, et al. Association between insomnia symptoms and functional status in U.S. older adults. J Gerontol B Psychol Sci Soc Sci. 2014;69 Suppl 1:S35–41.

6. Gao C, Chapagain NY, Scullin MK. Sleep Duration and Sleep Quality in Caregivers of Patients With Dementia: A Systematic Review and Meta-analysis. JAMA Netw Open. 2019;2(8):e199891.

7. Walsh CM, Ruoff L, Walker K, et al. Sleepless Night and Day, the Plight of Progressive Supranuclear Palsy. Sleep. 2017;40(11).

8. Dauphinot V, Faure R, Omrani S, et al. Exposure to anticholinergic and sedative drugs, risk of falls, and mortality: an elderly inpatient, multicenter cohort. J Clin Psychopharmacol. 2014;34(5):565–570.

9. Weich S, Pearce HL, Croft P, et al. Effect of anxiolytic and hypnotic drug prescriptions on mortality hazards: retrospective cohort study. BMJ. 2014;348:g1996.

10. Cipriani G, Lucetti C, Danti S, Nuti A. Sleep disturbances and dementia. Psychogeriatrics. 2015;15(1):65–74.

11. Thannickal TC, Moore RY, Nienhuis R, et al. Reduced number of hypocretin neurons in human narcolepsy. Neuron. 2000;27(3):469–474.

12. Oh J, Eser RA, Ehrenberg AJ, et al. Profound degeneration of wake-promoting neurons in Alzheimer’s disease. Alzheimers Dement. 2019.

13. Carpenter BD, Strauss M, Patterson MB. Sleep Disturbances in Community-Dwelling Patients with Alzheimer’s Disease. Clinical Gerontologist. 1996;16(2):35–49.

14. Moran M, Lynch CA, Walsh C, Coen R, Coakley D, Lawlor BA. Sleep disturbance in mild to moderate Alzheimer’s disease. Sleep Med. 2005;6(4):347–352.

15. Tractenberg RE, Singer CM, Kaye JA. Characterizing sleep problems in persons with Alzheimer’s disease and normal elderly. J Sleep Res. 2006;15(1):97–103.

16. Thannickal TC, Lai YY, Siegel JM. Hypocretin (orexin) cell loss in Parkinson’s disease. Brain. 2007;130(Pt 6):1586–1595.

17. Aziz A, Fronczek R, Maat-Schieman M, et al. Hypocretin and melanin-concentrating hormone in patients with Huntington disease. Brain Pathol. 2008;18(4):474–483.

18. Bittencourt JC. Anatomical organization of the melanin-concentrating hormone peptide family in the mammalian brain. Gen Comp Endocrinol. 2011;172(2):185–197.

19. Elias CF, Lee CE, Kelly JF, et al. Characterization of CART neurons in the rat and human hypothalamus. J Comp Neurol. 2001;432(1):1–19.

20. Croizier S, Cardot J, Brischoux F, Fellmann D, Griffond B, Risold PY. The vertebrate diencephalic MCH system: a versatile neuronal population in an evolving brain. Front Neuroendocrinol. 2013;34(2):65–87.

21. Hassani OK, Lee MG, Jones BE. Melanin-concentrating hormone neurons discharge in a reciprocal manner to orexin neurons across the sleep-wake cycle. Proc Natl Acad Sci U S A. 2009;106(7):2418–2422.

22. Jego S, Glasgow SD, Herrera CG, et al. Optogenetic identification of a rapid eye movement sleep modulatory circuit in the hypothalamus. Nat Neurosci. 2013;16(11):1637–1643.

23. Vetrivelan R, Kong D, Ferrari LL, et al. Melanin-concentrating hormone neurons specifically promote rapid eye movement sleep in mice. Neuroscience. 2016;336:102–113.

24. Qu D, Ludwig DS, Gammeltoft S, et al. A role for melanin-concentrating hormone in the central regulation of feeding behaviour. Nature. 1996;380(6571):243–247.

25. Diniz GB, Bittencourt JC. The Melanin-Concentrating Hormone as an Integrative Peptide Driving Motivated Behaviors. Front Syst Neurosci. 2017;11:32.

26. Burdakov D, Gerasimenko O, Verkhratsky A. Physiological changes in glucose differentially modulate the excitability of hypothalamic melanin-concentrating hormone and orexin neurons in situ. J Neurosci. 2005;25(9):2429–2433.

27. Barson JR, Morganstern I, Leibowitz SF. Complementary roles of orexin and melanin-concentrating hormone in feeding behavior. Int J Endocrinol. 2013;2013:983964.

28. Jiang H, Bruning JC. Melanin-Concentrating Hormone-Dependent Control of Feeding: When Volume Matters. Cell Metab. 2018;28(1):7–8.

29. Montine TJ, Phelps CH, Beach TG, et al. National Institute on Aging-Alzheimer’s Association guidelines for the neuropathologic assessment of Alzheimer’s disease: a practical approach. Acta Neuropathol. 2012;123(1):1–11.

30. Grinberg LT, Ferretti RE, Farfel JM, et al. Brain bank of the Brazilian aging brain study group - a milestone reached and more than 1,600 collected brains. Cell Tissue Bank. 2007;8(2):151–162.

31. Hyman BT, Phelps CH, Beach TG, et al. National Institute on Aging-Alzheimer’s Association guidelines for the neuropathologic assessment of Alzheimer’s disease. Alzheimers Dement. 2012;8(1):1–13.

32. Suemoto CK, Ferretti-Rebustini RE, Rodriguez RD, et al. Neuropathological diagnoses and clinical correlates in older adults in Brazil: A cross-sectional study. PLoS Med. 2017;14(3):e1002267.

33. Cairns NJ, Bigio EH, Mackenzie IR, et al. Neuropathologic diagnostic and nosologic criteria for frontotemporal lobar degeneration: consensus of the Consortium for Frontotemporal Lobar Degeneration. Acta Neuropathol. 2007;114(1):5–22.

34. Mai JK, Assheuer J, Paxinos G. Atlas of the Human Brain. 2. ed. San Diego, CA, USA: Elsevier Academic Press; 2004.

35. Ding SL, Royall JJ, Sunkin SM, et al. Comprehensive cellular-resolution atlas of the adult human brain. J Comp Neurol. 2016;524(16):3127–3481.

36. Swaab DF. Chapter 14 Lateral hypothalamic area (LHA), including the perifornical area and intermediate hypothalamic area (IHA). Handb Clin Neurol. 2003;79:281–284.

37. Heinsen H, Arzberger T, Schmitz C. Celloidin mounting (embedding without infiltration) - a new, simple and reliable method for producing serial sections of high thickness through complete human brains and its application to stereological and immunohistochemical investigations. J Chem Neuroanat. 2000;20(1):49–59.

38. Ehrenberg AJ, Nguy AK, Theofilas P, et al. Quantifying the accretion of hyperphosphorylated tau in the locus coeruleus and dorsal raphe nucleus: the pathological building blocks of early Alzheimer’s disease. Neuropathol Appl Neurobiol. 2017;43(5):393–408.

39. West MJ, Slomianka L, Gundersen HJ. Unbiased stereological estimation of the total number of neurons in thesubdivisions of the rat hippocampus using the optical fractionator. Anat Rec. 1991;231(4):482–497.

40. Gundersen HJ, Jensen EB, Kieu K, Nielsen J. The efficiency of systematic sampling in stereology--reconsidered. J Microsc. 1999;193(Pt 3):199–211.

41. Schmitz C, Hof PR. Design-based stereology in neuroscience. Neuroscience. 2005;130(4):813–831.

42. Diniz GB, Bittencourt JC. The Melanin-Concentrating Hormone (MCH) System: A Tale of Two Peptides. Front Neurosci. 2019;13:1280.

43. Arrigoni E, Chee MJS, Fuller PM. To eat or to sleep: That is a lateral hypothalamic question. Neuropharmacology. 2019;154:34–49.

44. Walsh CM, Ruoff L, Varbel J, et al. Rest-activity rhythm disruption in progressive supranuclear palsy. Sleep Med. 2016;22:50–56.

45. Montplaisir J, Petit D, Decary A, et al. Sleep and quantitative EEG in patients with progressive supranuclear palsy. Neurology. 1997;49(4):999–1003.

46. Peyron C, Tighe DK, van den Pol AN, et al. Neurons containing hypocretin (orexin) project to multiple neuronal systems. J Neurosci. 1998;18(23):9996–10015.

47. Elias CF, Saper CB, Maratos-Flier E, et al. Chemically defined projections linking the mediobasal hypothalamus and the lateral hypothalamic area. J Comp Neurol. 1998;402(4):442–459.

48. Schmidt FM, Kratzsch J, Gertz HJ, et al. Cerebrospinal fluid melanin-concentrating hormone (MCH) and hypocretin-1 (HCRT-1, orexin-A) in Alzheimer’s disease. PLoS One. 2013;8(5):e63136.

49. Liguori C, Romigi A, Nuccetelli M, et al. Orexinergic system dysregulation, sleep impairment, and cognitive decline in Alzheimer disease. JAMA Neurol. 2014;71(12):1498–1505.

50. Raskind MA, Peskind ER, Halter JB, Jimerson DC. Norepinephrine and MHPG levels in CSF and plasma in Alzheimer’s disease. Arch Gen Psychiatry. 1984;41(4):343–346.

51. Apergis-Schoute J, Iordanidou P, Faure C, et al. Optogenetic evidence for inhibitory signaling from orexin to MCH neurons via local microcircuits. J Neurosci. 2015;35(14):5435–5441.

52. Tsunematsu T, Ueno T, Tabuchi S, et al. Optogenetic manipulation of activity and temporally controlled cell-specific ablation reveal a role for MCH neurons in sleep/wake regulation. J Neurosci. 2014;34(20):6896–6909.

53. Varin C, Luppi PH, Fort P. Melanin-concentrating hormone-expressing neurons adjust slow-wave sleep dynamics to catalyze paradoxical (REM) sleep. Sleep. 2018;41(6).

54. Lucey BP, McCullough A, Landsness EC, et al. Reduced non-rapid eye movement sleep is associated with tau pathology in early Alzheimer’s disease. Sci Transl Med. 2019;11(474).

